# Deep Learning for Rare Disease: A Scoping Review

**DOI:** 10.1101/2022.06.29.22277046

**Authors:** Junghwan Lee, Cong Liu, Junyoung Kim, Zhehuan Chen, Yingcheng Sun, James R. Rogers, Wendy K. Chung, Chunhua Weng

## Abstract

Although individually rare, collectively more than 7,000 rare diseases affect about 10% of patients. Each of the rare diseases impacts the quality of life for patients and their families, and incurs significant societal costs. The low prevalence of each rare disease causes formidable challenges in accurately diagnosing and caring for these patients and engaging participants in research to advance treatments. Deep learning has advanced many scientific fields and has been applied to many healthcare tasks. This study reviewed the current uses of deep learning to advance rare disease research. Among the 332 reviewed articles, we found that deep learning has been actively used for rare neoplastic diseases (250/332), followed by rare genetic diseases (170/332) and rare neurological diseases (127/332). Convolutional neural networks (307/332) were the most frequently used deep learning architecture, presumably because image data were the most commonly available data type in rare disease research. Diagnosis is the main focus of rare disease research using deep learning (263/332). We summarized the challenges and future research directions for leveraging deep learning to advance rare disease research.

## Introduction

Currently about 7,000 rare diseases (the diseases each affecting 1/2,000 patients in the European Union or fewer than 200,000 people in the United States) affect over 350 million patients worldwide[1]. Many of these rare diseases significantly decrease the quality of life for those affected and are associated with substantial healthcare costs [2]. Thus, there is a tremendous need to provide care for rare disease patients. However, there are significant challenges in delivering high-quality care to rare disease patients. First, timely and accurate diagnosis remain difficult and rare disease patients spend an average of 6 years seeking an accurate diagnosis from the onset of symptoms[3]. Second, research on pathophysiological mechanisms to understand these rare diseases and to develop treatments is limited. Both challenges are mainly due to the low prevalence of individual rare disease, insufficient numbers of rare disease specialists with adequate disease-specific knowledge, lack of access to those specialists, and insufficient resources and infrastructure for rare disease research[4].

An emerging method to overcome some of these aforementioned challenges is to provide computer-based aid for user augmentation in the clinical workflow. As a type of machine learning where models contain multiple layers of neural networks[5], deep learning has advanced many scientific fields such as computer vision, speech recognition and natural language processing[5]. It has also been applied to various healthcare tasks[6, 7], including disease diagnosis[8, 9], disease onset prediction[10], mortality prediction[11], length of stay in hospital prediction[12], phenotyping[13], and novel drug discovery[14]. Given these promising applications, there has been great interest in using deep learning to address the challenges for rare diseases.

Despite its emerging interests, there is no comprehensive overview of rare disease research using deep learning. Schaefer et al.[4] recently conducted a review on the use of general machine learning for rare diseases. In this study, we conducted a comprehensive literature review of deep learning for rare diseases and accompanying opportunities and challenges.

## Methods

This scoping review was conducted by following the Preferred Reporting Items for Systematic reviews and Meta-Analyses extension for scoping reviews (PRISMA-ScR) guideline[15]. This study does not include any materials or experiments that need ethical approval.

### Literature search

We systematically searched for candidate articles in four databases: 1) PubMed; 2) the Association of Computing Machinery (ACM) Digital Library; 3) the Institute of Electrical and Electronics Engineers (IEEE) Xplore; and 4) DBLP. The following search criteria were used: (1) written in English; (2) published or publicly available (e.g., conference proceeding) between January 1, 2010 and December 31, 2021; 3) topic of interest includes deep learning and rare disease.

The general search string was constructed by combining terms representing deep learning (“machine learning” or “deep learning”) and “rare disease”. For DBLP, we only used the term “rare disease” due to the limitation of advanced search features of the database. For PubMed that indexes a broad range of scientific literature on life sciences and biomedical topics, we used 7,138 individual rare disease names with the terms “deep learning” and “machine learning”. Those 7,138 rare disease names were obtained from Orphanet[16], one of the largest knowledge databases for rare diseases. 7,138 individual rare disease names are available in **Supplementary Table 1**. The initial search was conducted in June 2021. An additional search to collect articles published in 2021 was conducted in May 2022. **Table 1** shows the search strings for each database.

**Table 1.** Search strings for four databases.

| Database | Search strings | The number of search strings |
| --- | --- | --- |
| Association of Computing Machinery (ACM) Digital Library | (“machine learning” AND “rare disease”)<br>(“deep learning” AND “rare disease”) | 2 |
| Institute of Electrical and Electronics Engineers (IEEE) Xplore | (“machine learning” AND “rare disease”)<br>(“deep learning” AND “rare disease”) | 2 |
| PubMed | (“machine learning” AND one of 7,138 individual rare disease names),<br>(“deep learning” AND one of 7,138 individual rare disease names) | 14,276 |
| DBLP | “rare disease” | 1 |

### Literature selection

After candidate articles were obtained, we conducted abstract and full-text screening. Exclusion criteria were: 1) not published in a peer-reviewed journal or conference proceeding (e.g., preprint); 2) not original research (e.g., editorial); 3) reviews; 4) not human subject research; and 5) topic of interest does not include deep learning. We excluded studies that solely used multilayer perceptron (MLP) as their deep learning architectures since MLP alone does not represent the state-of-the-art deep learning architectures; and 6) topic of interest is not on rare diseases. All abstracts and full-texts were initially screened by one reviewer (JL), followed by the verifications done by three independent reviewers (JK, ZC and YS). Disagreements were discussed among the reviewers until consensus was reached. Covidence[17] was used for citation and screening management.

### Data extraction and synthesis

One reviewer (JL) developed and extracted metadata for each article. Three reviewers (JK, ZC and YS) verified and refined the metadata for completeness. The resulting metadata include 1) publication year; 2) analytic task; 3) data type; 4) data size; 5) purpose of using deep learning; 6) deep learning architecture; 7) rare disease; 8) rare disease group; and 9) challenges in using deep learning for rare diseases. **Table 2** summarizes the metadata elements.

**Table 2.** Metadata extracted from the articles.

| Metadata | Definition | Categories or Range |
| --- | --- | --- |
| Publication year | Year the article was published (or deposited if conference proceeding). | Year between 2010-2021 |
| Analytic task | Goal of the article using deep learning for rare disease; classified as at least one of the following. | -Diagnosis: to accurately identify the rare disease |
|  |  | -Prognosis: to forecast the course or outcome of the rare disease |
|  |  | -Treatment: to predict how well an interventional regimen will perform in addressing the rare disease |
|  |  | -Characterization: to investigate characteristics or mechanisms of the rare disease |
|  |  | -Others: if the model goal cannot be characterized in one of the above categories |
| Data type | Where the underlying inputs for deep learning model(s) are derived from; classified as at least one of the following. | -Image data: data produced by scanning objects with an optical or electronic devices (e.g., Computed tomography (CT) scan, Magnetic resonance imaging (MRI), Fundus images) |
|  |  | -Measurement data: data produced by measuring biological or physiological objects (e.g., Omics data, Lab test data, Electroencephalography (EEG), Electromyography (EMG)) |
|  |  | -Encounter data: longitudinal data obtained from recorded patients' encounters (e.g., Billing codes, Medication records, Clinical notes) |
|  |  | -Literature data: data obtained from scientific literature (e.g., published articles) |
| Data size | Quantity tied to a specific unit of input that makes up a particular data type sample. The data size for each data type was measured based on the smallest unit of input for a model before any data augmentation was applied. | -Image data: # of sliced images; # of images |
|  |  | -Measurement data: # of biospecimens; # of lab tests |
|  |  | -Encounter data: # of individuals |
|  |  | -Literature data: # of articles |
| Purpose of using deep learning | Purpose of using deep learning for rare disease identified from the articles; classified as at least one of the categories. | -Proof-of-concept |
|  |  | -To improve performance of the analytic task |
|  |  | -To reduce manual effort |
|  |  | -To address lack of data |
| Deep learning architecture | Deep learning architecture used to achieve the goal of the articles; classified as at least one of the categories. | -Autoencoders (AE) |
|  |  | -Convolutional neural networks (CNN) |
|  |  | -Generative adversarial networks (GAN) |
|  |  | -Recurrent neural networks (RNN) |
|  |  | -Self-attention-based architectures |
|  |  | -Unsupervised embedding |
| Rare disease | Condition(s) studied within the article; identification of a rare disease is based on available Orphanet identifier (e.g., Huntington disease OPRHA:399). If an article aims to investigate specific characteristics of rare diseases that can be applied to general rare diseases (e.g., a method to learn from scarce data), we denoted the article studied “General” rare disease. |  |
| Rare disease group | Classification(s) of the rare disease(s) studied within the article; classification is based on highest available Orphanet ancestor identifier (e.g., Rare neurologic diseases OPRHA:98006). |  |
| Challenges in using deep learning for rare diseases | Challenges in using deep learning for rare disease and future directions to improve that were identified by the authors of the articles; classified as at least one of the following. | -Insufficient data: insufficient data amounts or data types to fully utilize deep learning models |
|  |  | -Data quality issue: biases, class imbalance, confounding factors in the data |
|  |  | -Limited choice of evaluation: further evaluations are required for more robust evaluation |
|  |  | -Lack of external validation: lack of validation of the results using external data |
|  |  | -Limited choice of models: validation by using other deep learning architectures is required |
|  |  | -Difficulties in training: the amount of resource (e.g., training time, computational power) to train deep learning models exceeds what is available for real-world application; issues in training deep learning models (e.g., overfitting) |
|  |  | -Lack of interpretability: deep learning models are not interpretable |
|  |  | -Lack of generalizability: lack of generalizability to other rare diseases |

## Results

Figure 1. displays the Preferred Reporting Items for Systematic reviews and Meta-Analyses (PRISMA) flow diagram for article selection. Of 4,516 articles found after de-duplication, 812 articles were included in full-text screening. After full-text screening, 332 articles were included for the final analysis. The comprehensive list of all 332 articles with their metadata elements extracted is available in **Supplementary Table 2**.

**Figure 1.**
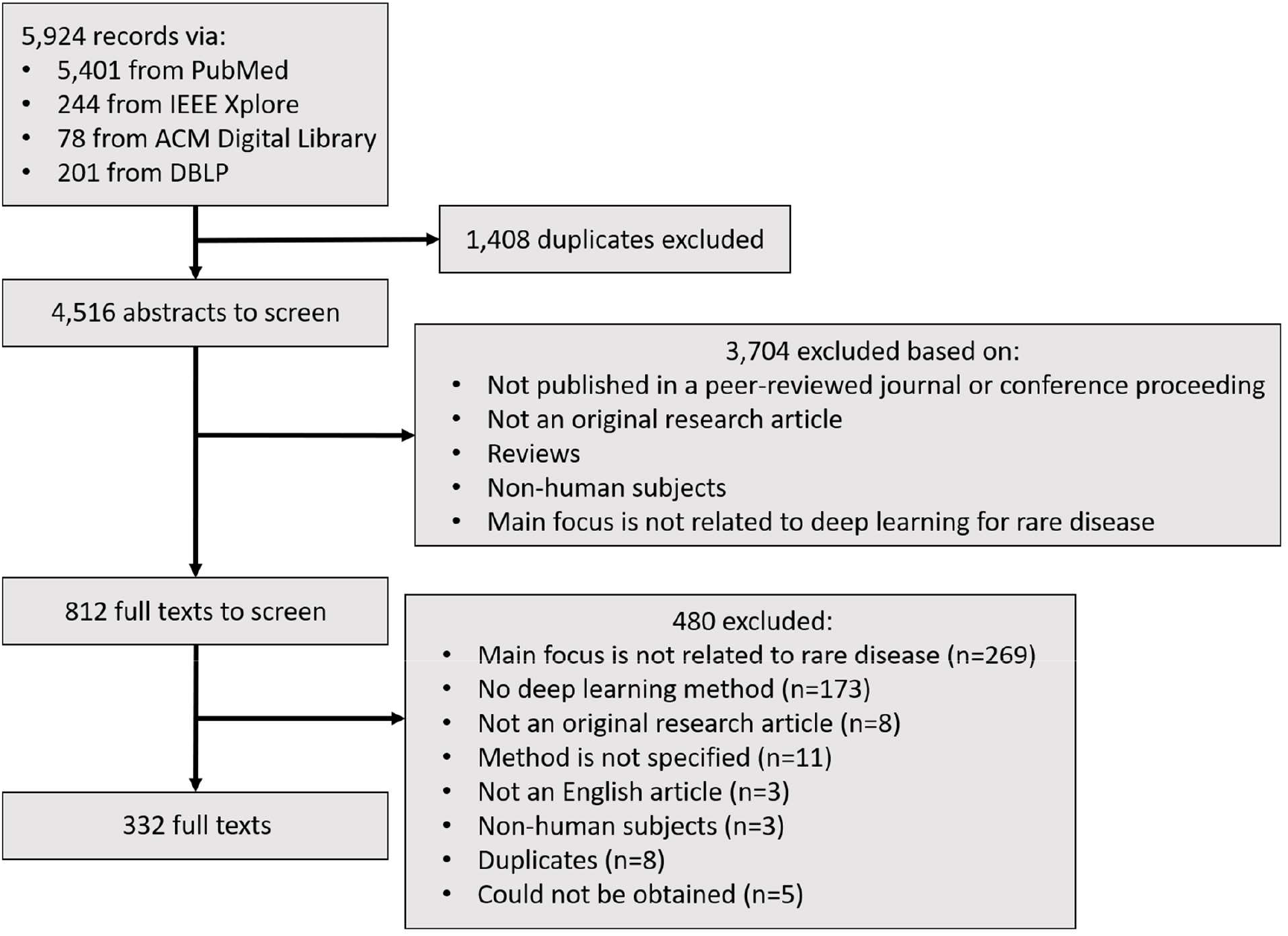
PRISMA flow diagram for literature selection.

### Yearly publication trend

**Table 3** shows the number of yearly publications in deep learning for rare disease, deep learning for any disease, and deep learning in general. The number of deep learning for any disease publications was estimated by search using the keyword “deep learning” and “disease” in Google Scholar. The number of deep learning publications was estimated by search using the keyword “deep learning” in Google Scholar. The number of publications in deep learning for rare disease was zero before 2017, but increased sharply from 10 article in 2017 to 145 articles in 2021. The number of publications in deep learning for rare disease and deep learning for any disease both showed greater growth trend than the number of publications in deep learning.

**Table 3.** The yearly number of publications in deep learning for rare disease, deep learning for any disease, and deep learning, respectively.

| Counts | 2010 | 2011 | 2012 | 2013 | 2014 | 2015 | 2016 | 2017 | 2018 | 2019 | 2020 | 2021 |
| --- | --- | --- | --- | --- | --- | --- | --- | --- | --- | --- | --- | --- |
| # of publications in deep learning for rare disease | 0 | 0 | 0 | 0 | 0 | 0 | 0 | 10 | 31 | 40 | 106 | 145 |
| # of publications in deep learning for any disease | 795 | 866 | 921 | 1090 | 1360 | 2030 | 3820 | 7720 | 16000 | 31000 | 45800 | 50200 |
| # of publications in deep learning | 5470 | 6010 | 7270 | 8490 | 11500 | 18500 | 42200 | 97900 | 206000 | 282000 | 268000 | 149000 |

### Analytic task for rare disease

20 articles focused on multiple analytic tasks and thus qualified for multiple categories. Of the 332 articles, 263 articles (79.2%) focused on diagnosis, 43 (13.0%) on prognosis, 29 (8.7%) on treatment, and 8 on characterization (2.4%) (**Figure 2A**). Diagnosis was the dominant focus of rare disease research using deep learning.

**Figure 2.**
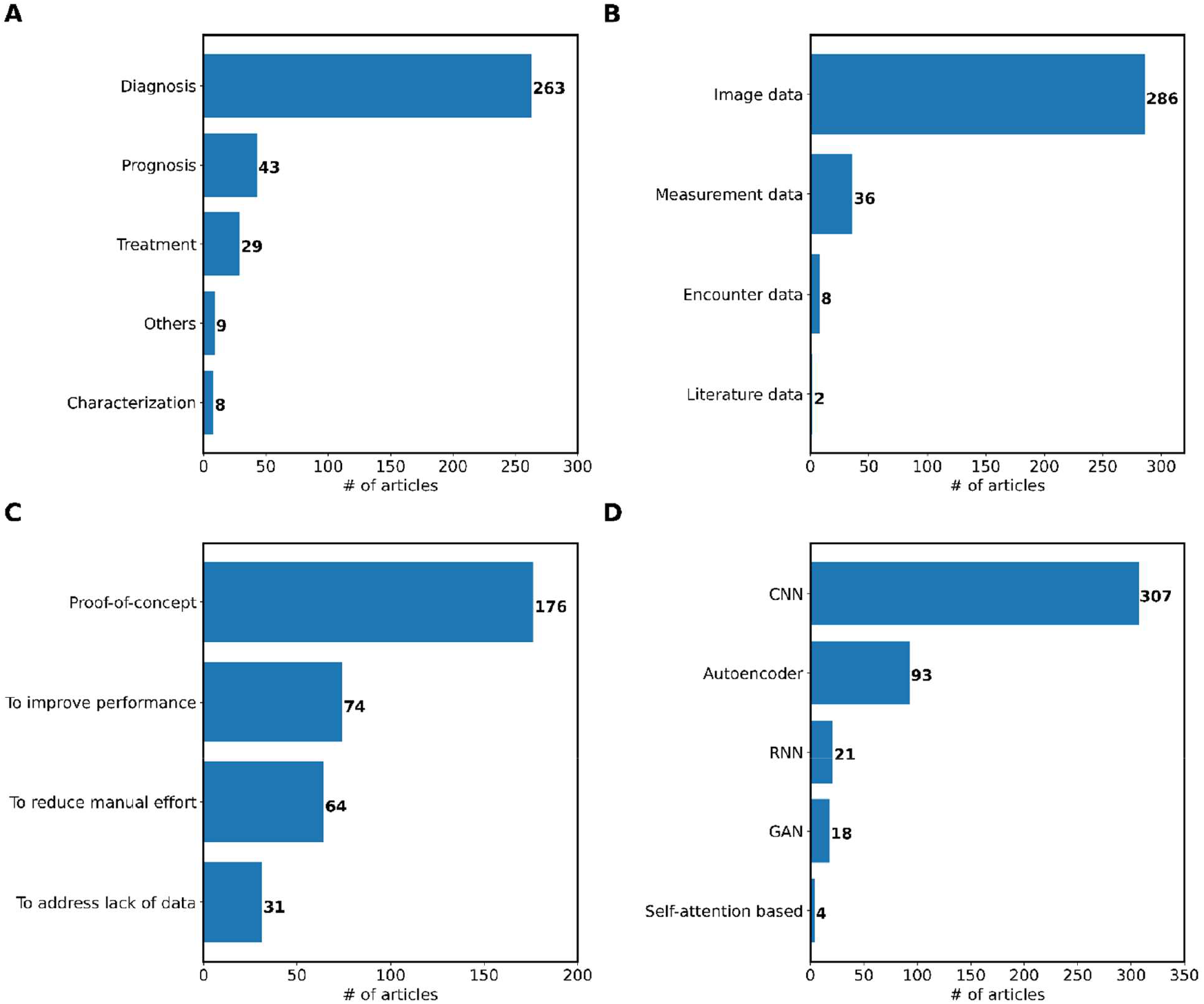
The number of articles identified for each (**A**) analytic task for rare disease, (**B**) data type, (**C**) purpose of using deep learning, and (**D**) deep learning architecture.

### Data types and sizes

As shown in **Figure 2B**, image data were the predominant data type used in 286 (86.1%) articles, while measurement data, encounter data, and literature data were used in 36 (10.8%), 8 (2.4%), and 2 (0.6%) articles, respectively. Specifically, MRI (85 articles) and CT (70 articles) were frequently used among image data (**Supplementary Table 2**).

We measured the data size for each data type based on the number of instances before any data augmentation was applied. An instance is defined as a single input for the deep learning model in the study. For example, we measured the number of patches instead of the number of images if the input of a deep learning model was patches of an image. The entire data size was measured before any split was conducted (e.g., split into training, validation, and test set). If multiple datasets were used for separate experiments independently, we measured the size of the largest dataset. Articles that did not clarify the size of data were excluded from the analysis. The median data sizes for image, encounter, and measurement data are 2,762 (interquartile range [IQR]: 790.5 – 12,817), 193,676 (IQR: 49,727.5 – 2,090,819), and 1,358 (IQR: 498 – 10,148) (**Figure 3**). We did not calculate median and IQR of the data size for literature data since it was used by only 2 articles.

**Figure 3.**
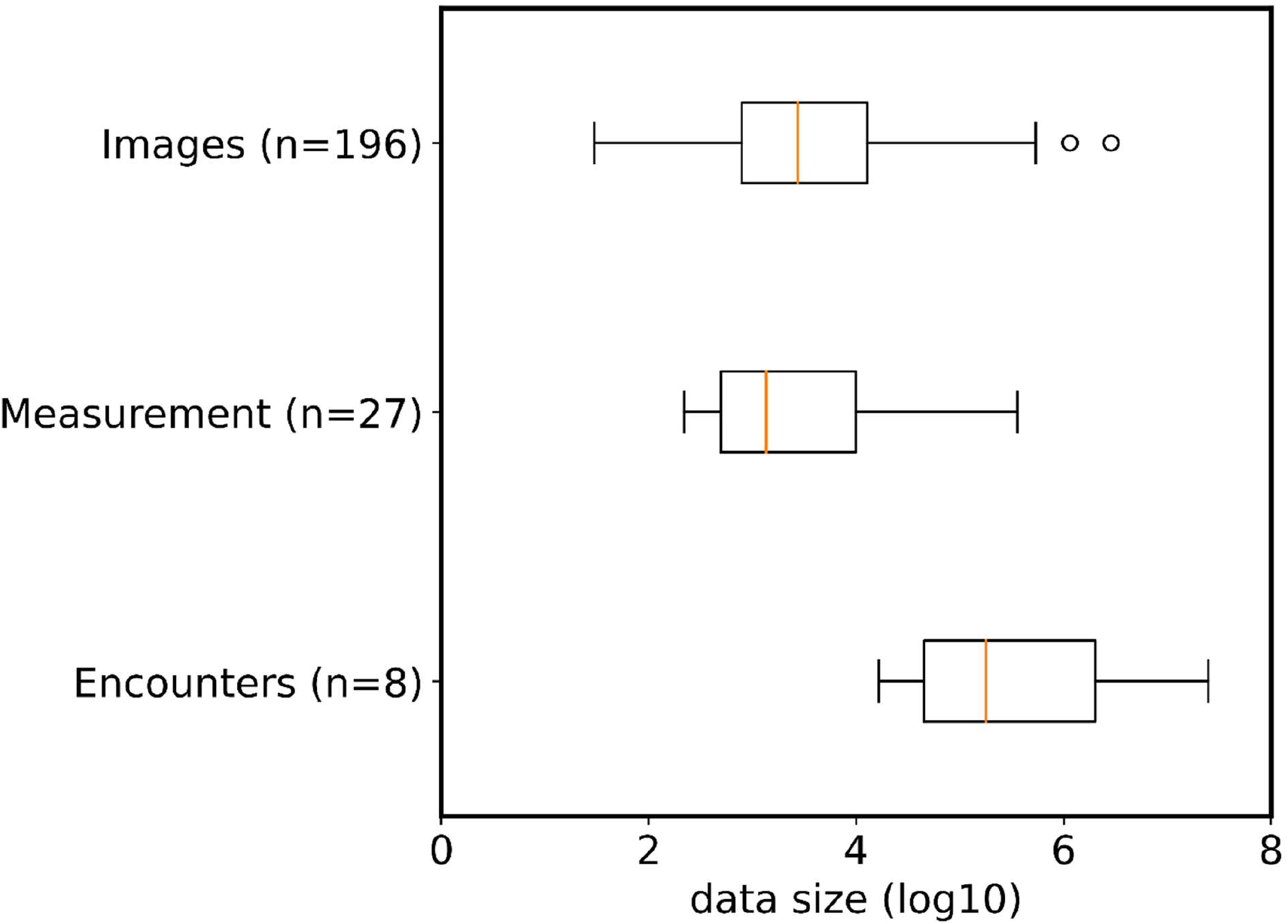
Box plots of data size for all data types.

We found that publicly available datasets were frequently used. Commonly used datasets included Brain Tumor Segmentation Challenge (BraTS) dataset[18], MIMIC-III[19] that contains various encounters from electronic health records obtained from the patients in critical care, and The Cancer Genome Atlas (TCGA) data portal[20] where genomic data of various cancers are available.

### Purpose of using deep learning

We classified the reviewed articles based on the purpose of using deep learning: why the study used deep learning over other traditional statistical or machine learning methods? **Figure 2C** shows the number of articles identified for each purpose of using deep learning. 13 articles had two purposes and hence were included in both categories. 176 articles (53.0%) simply used deep learning based on the successful application in other domains or in similar tasks with other rare disease studies without specifying the purpose. The second most common purpose of using deep learning was to improve the performance of the analytic task (74 articles [22.3%]). Deep learning was used in 64 articles (19.3%) to reduce manual effort for their target analytic tasks, specifically to reduce manual feature engineering effort or automation of target analytic tasks. 31 articles (9.3%) utilized deep learning to mitigate lack of data issue.

### Deep learning architecture

We recognized a deep learning models used in the articles include combinations of different architectures. **Figure 2D** shows the number of articles using each deep learning architecture to build their deep learning models: convolutional neural networks (CNN; 307 articles [92.5%]); autoencoders (AE; 93 articles [28.0%]); recurrent neural networks (RNN; 21 articles [6.3%]); generative adversarial networks (GAN; 18 articles [5.4%]); and self-attention-based architecture (4 articles [1.2%]). **Figure 4** visually illustrates the deep learning architectures. 100 articles (30.1%) used more than one deep learning architectures. We discussed the uses of each deep learning architecture with selected representative articles in the following subsections. Details of the architecture for all reviewed articles are available in **Supplementary Table 2**.

**Figure 4.**
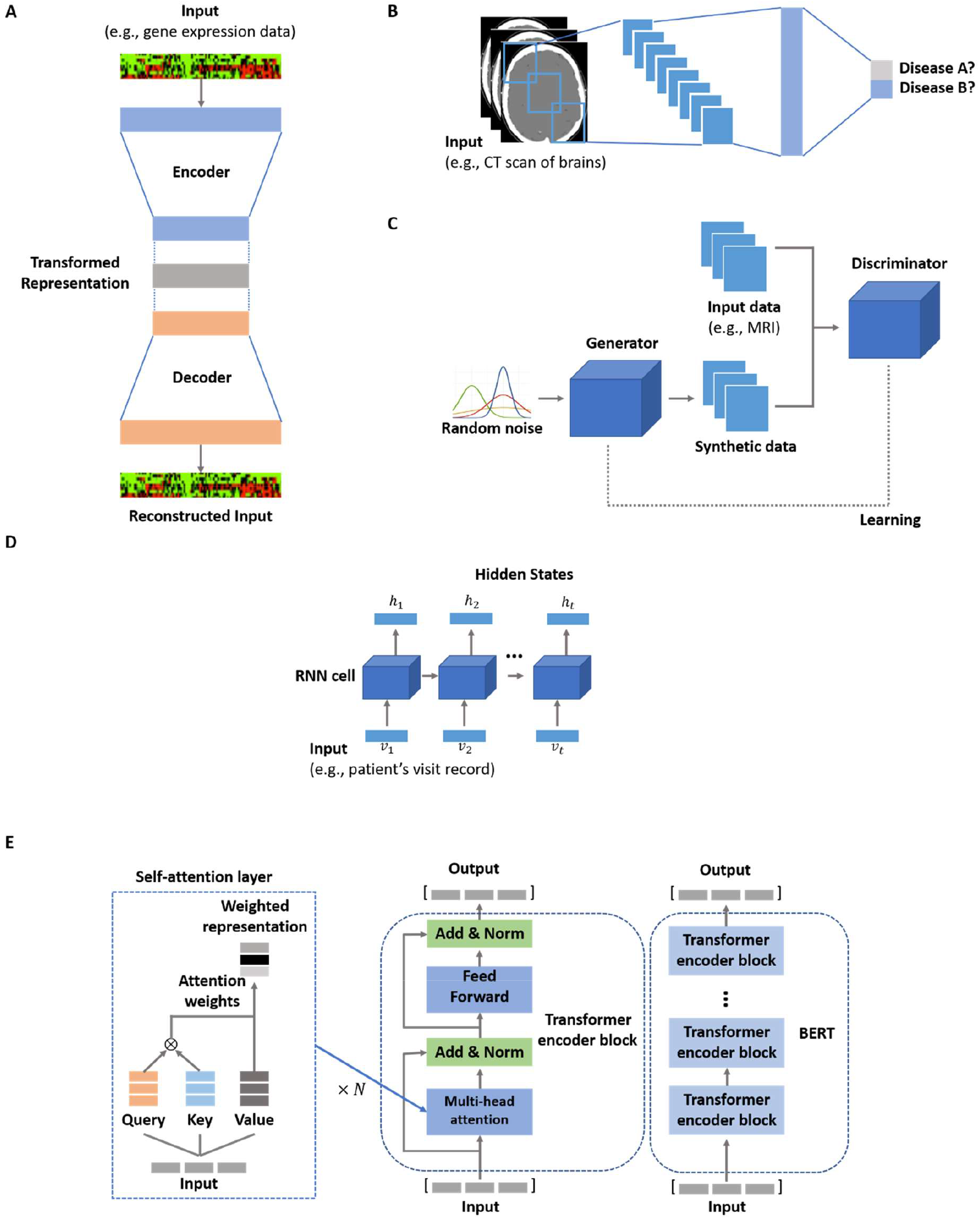
Visual illustrations of deep learning architectures. (**A**) Autoencoder (AE). (**B**) Convolutional neural networks (CNN). (**C**) Generative adversarial networks (GAN). (**D**) Recurrent neural networks (RNN). (**E**) Transformer encoder and BERT.

### Autoencoders (AE)

Autoencoder (AE) is a neural network architecture that is trained to reconstruct a given input with internal hidden layers consisting of two parts: encoder and decoder[21]. Encoder transforms an input data into a representation in a distributed space and decoder reconstructs the input from the transformed representation (**Figure 4A**), which learns useful information of the input data in an efficient way, generally having low dimensionality. Image data (83 articles) were frequently used with AEs (**Figure 5A**). The most common analytic task is diagnosis (**Figure 5B**).

**Figure 5.**
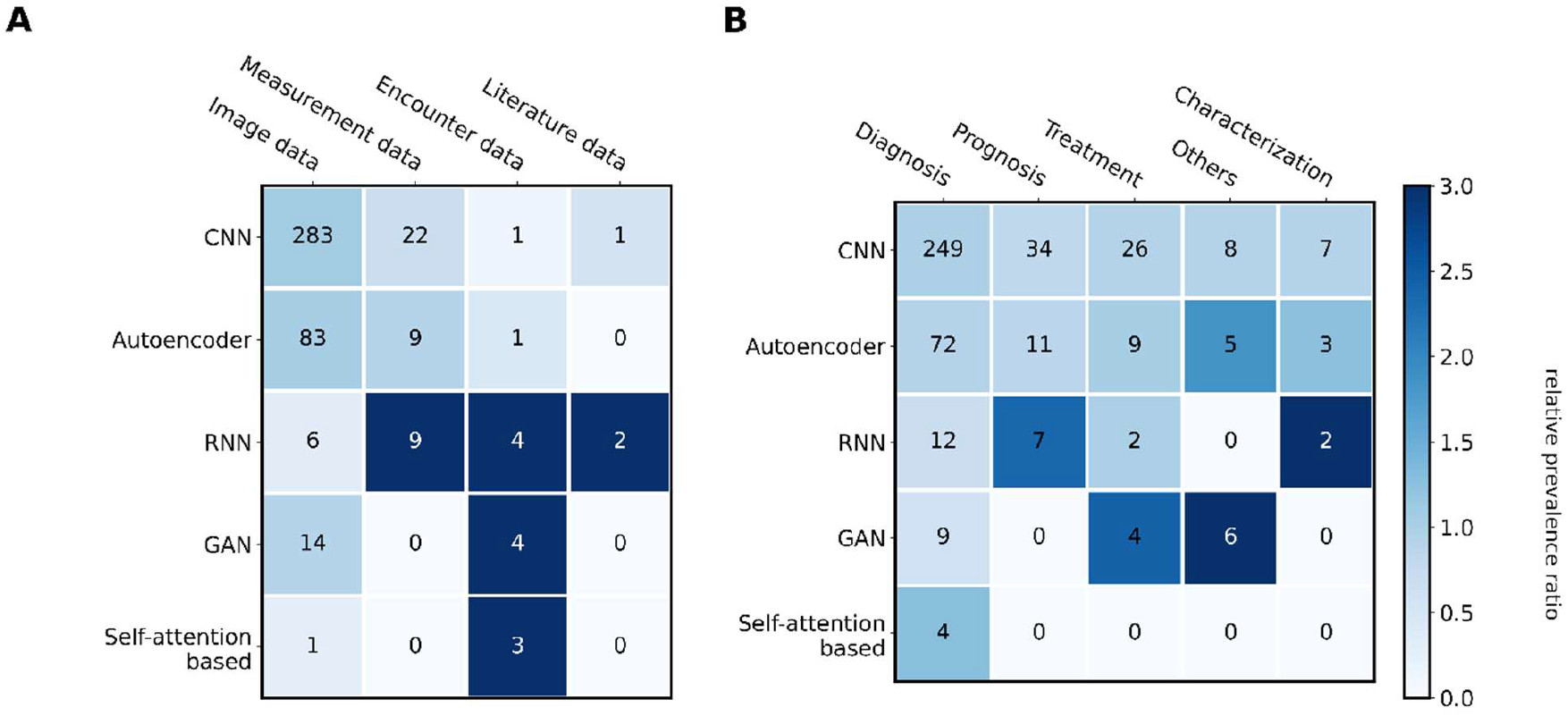
(**A**) A heatmap of deep learning architecture vs. data type and (**B**) deep learning architecture vs. analytic task. The number in each cell in the heatmap represents the number of articles that used the corresponding deep learning architecture and data type in (**A**) and the number of articles that used the corresponding deep learning architecture and analytic task in (**B**). The color of each cell in the heatmap represents the relative prevalence ratio (RPR) based on deep learning architecture. RPR for each cell was calculated as dividing the prevalence of the metadata element pair based on deep learning architecture (the number in a cell / row sum) by the baseline prevalence of the deep learning architecture. The baseline prevalence of each deep learning architecture is calculated as the number of articles identified with each deep learning architecture divided by the total number of articles. RPR was clipped by maximum value=3.0 for visualization.

The transformed representation by using AEs was often directly used for target analytic tasks with supervision[22-26] or used for clustering of the data[27-29]. For example, [22] used transformed representation by encoder to predict survival of Neuroblastoma patients. In [27, 28], omics data were encoded by AEs and then transformed representations were used to cluster tuberculosis patient and subtypes of neuroblastoma respectively. U-Net[30] and its variants (e.g., V-Net[31]), which have encoder-decoder architecture combined with convolutional neural network, are predominantly chosen architecture in the most of image segmentation tasks for rare disease. For example, [32] used U-Net for segmentation of cardiac magnetic resonance imaging to predict prognosis of patients with tetralogy of Fallot.

Variational autoencoder (VAE)[33] has characteristics of generative models and autoencoders and was used to learn efficient representation of input since the encoding of the input is restricted by prior distribution. For example, [34] used VAE to learn efficient encoding of input data for status prediction of glioblastoma patients and [35] used VAE with convolutional neural networks to cluster atomistic molecular dynamics for Gaucher disease research.

### Convolutional neural networks (CNN)

CNNs are neural networks designed for data having grid-like topological features (**Figure 4B**). CNNs’ impressive capability to learn local conjunctions of features has shown significant success in many practical applications[5]. Image data (283 articles) were predominantly used with CNNs, followed by measurement data (21 articles) (**Figure 5A**).

Various state-of-the-art CNN architectures were used in the articles. Commonly used CNN architectures includes but not limited to VGGNet[36], GoogLeNet[37], ResNet[38], Inception-V3[39], and DenseNet[40]. A few studies proposed novel architectures based on the existing CNN architectures for their target rare diseases and analytic tasks. For example, [41] proposed a novel architecture for tuberculosis detection based on chest X-ray using ResNet and attention; [42] proposed a novel architecture for retinitis pigmentosa detection based on optical coherence tomography using the idea of ResNet and DenseNet.

DeepMedic[43], a CNN architecture especially developed for 3D brain MRI, is one of the widely used CNN architectures for image segmentation tasks for rare brain tumors. For example, DeepMedic was used to diagnose glioblastoma[44-46], primary central nervous system lymphoma[47], and meningioma[48, 49]. As mentioned above, state-of-the-art CNN architectures combined with encoder-decoder architecture (e.g., U-Net[30]) are also widely used in image segmentation tasks for rare disease.

Transfer learning was a common strategy to solve insufficient data problem for CNNs. In the studies that used CNNs, CNNs were often pre-trained with publicly available datasets such as Imagenet[50] or MS-COCO[51], then pre-trained CNNs were transferred to be used for target analytic tasks. In several studies, datasets that are more closely related to their target analytic tasks were used. For example, [45, 47, 48, 52] used brain MRI dataset to pre-train CNNs for their brain MRI segmentation task. [53] reported improved performance by using CNNs pre-trained with task-related datasets than CNNs pre-trained with Imagenet.

CNNs were used with other data types by utilizing CNN’s capacity to learn patterns in data such as genomic data[54, 55], EMG signals[56], electroencephalography[57], protein-protein molecular dynamics[35], and published rare disease literature[58]. For example, [54] applied CNN to SNP maps to predict occurrences of Down syndrome. [56] converted EMG signals to 2-dimensional time-frequency representation and applied CNN to classify amyotrophic lateral sclerosis. [58] used CNN to find the relationship between disabilities and rare diseases in published rare disease literature.

CNNs were also used to learn representations of the nodes in graph data. For example, [59] used graph convolutional networks (GCN)[60] to learn the representations of rare disease phenotypes from disease ontology graph where phenotypes are connected nodes; [61] applied GCN to antiplasmodial hit compounds dataset for discovery of antimalarial drugs; and [62] used GCN to learn representations of diseases using disease knowledge graph in generating synthetic radiology report for rare disease patients.

### Generative adversarial networks (GAN)

GANs are generative models that can generate synthetic data through an adversarial training of generator and discriminator in the models[63]. The generator is trained to generate realistic input-like data, while the discriminator is trained to distinguish between the generated data and real input data (**Figure 4C**). GANs were only used with Image data (14 articles) and encounter data (4 articles) (**Figure 5A**). While 50% of the studies used GANs aimed to diagnosis tasks, 6 articles, which are classified as others, purely focused on synthetic data generation without specifying analytic task (**Figure 5B**).

GANs were often used to augment data for training. This is especially useful when there was lack of data or supervision (i.e., labels) for training a model, which is important for rare disease research that often suffers from lack of data. For example, [62, 64] used GAN to generate synthetic X-ray images and encounter visits of rare disease patients. In [65-67], GANs were used to learn efficient representation from unlabeled data in semi-supervised setting (i.e., lack of supervision) to detect rare disease patients. [68] aimed to generate realistic synthetic CT images based on magnetic resonance images using CNN-based GAN. Besides utilizing GANs for generating synthetic data, the adversarial training scheme has also been used to improve the performance of the model for an analytic task[69-71].

### Recurrent neural networks (RNN)

RNNs are a family of neural networks for modeling sequential data [21]. RNNs consist of RNN cell that is specially designed to process input sequence at each sequential timestamp (**Figure 4D**). Among many RNN cell variants, Long Short-Term Memory (LSTM)[72] was the most frequently used in the articles that used RNNs. Measurement data and encounter data were more frequently used with RNNs (**Figure 5A**). While the most common analytic task is diagnosis, prognosis and characterization were more frequently involved with RNNs (**Figure 5B**).

Accumulation of medical history for accurate diagnosis of rare diseases often takes several years. RNNs are capable of learning long-term dependencies and can be suitable architectures for diagnosis and prognosis of rare diseases[64, 66, 67, 73]. For example, [73] used RNN to learn from longitudinal billing codes for mortality prediction of rare disease. Likewise, longitudinal measurement was also found to be a promising data type to utilize the RNNs[74-76]. Sequential images of a rare disease patient (e.g., MRI) were applied to RNNs for prognosis of the disease[77, 78]. Multiple slices or pixels of a single image of a rare disease patient were applied to RNNs to detect the region affected by the disease[71, 79].

Bidirectional RNNs often outperform unidirectional RNNs by utilizing two directional RNNs instead of the single direction of left-to-right RNNs[80]. There were a few studies that used bidirectional RNNs: [78] used bidirectional RNN for the prognosis of glioblastoma patients; [58] used bidirectional RNN to identify the relationships between disabilities and diseases from rare disease corpus; and [81] used CNNs and bidirectional RNN to classify medulloblastoma.

### Self-attention-based architectures

Self-attention-based architectures utilize attention-weighted features where attention weights are calculated based on the relevance to the downstream task and have gained popularity due to its impressive performance in many application domains. Transformer[82] and BERT[83], the two most widely used self-attention-based architectures, were used in the articles (**Figure 4E**). The articles used self-attention-based architectures only focused on diagnosis task (**Figure 5A**). Encounter data were predominantly used with self-attention-based architectures (**Figure 5B**).

[67] used Transformer to learn representations of conditions and drugs recorded in visits of patients for identifying rare disease patients. [84] used Transformer to generate radiology report for rare diseases in data-scarce setting (i.e., few shot learning). [85, 86] used BERT to identify rare disease patients with encounter data.

### Rare diseases and rare disease groups

83 unique rare diseases were identified from the reviewed articles. **Table 4** shows the 10 most frequently studied rare diseases. Multiple rare diseases were often studied in one article. We classified the rare diseases identified from the reviewed articles into rare disease groups using the hierarchical structure defined by Orphanet. A rare disease can qualify for multiple groups and resulted in the 83 unique rare diseases being classified into 30 rare disease groups. **Figure 6A** shows the 10 most frequently studied rare disease groups in the reviewed articles, with the three most common being rare neoplastic diseases (75.3%), rare genetic diseases (51.2%) and rare neurological diseases (38.3%), respectively. **Figure 6B** shows the number of articles identified for each challenge. The full list of rare disease groups with the count of articles for each rare disease group is available in **Supplementary Table 3**.

**Table 4.** The 10 most frequently studied rare diseases. The estimated prevalence in the worldwide population of rare diseases were obtained from Orphanet.

| Disease | Number of articles<br>(frequency) | Estimated prevalence in<br>the general population |
| --- | --- | --- |
| Tuberculosis | 41 (12.3%) | 1-5 / 10,000 |
| Glioblastoma | 38 (11.4%) | 1-9 / 100,000 |
| Nasopharyngeal carcinoma | 34 (10.2%) | 1-9 / 100,000 |
| Malaria | 27 (8.1%) | 1-9 / 100,000 |
| Central serous chorioretinopathy | 10 (3.0%) | - |
| Retinopathy of prematurity | 10 (3.0%) | 1-5 / 10,000 |
| Meningioma | 9 (2.7%) | - |
| Retinitis pigmentosa | 7 (2.1%) | 1-5 / 10,000 |
| Amyotrophic lateral sclerosis | 7 (2.1%) | 1-9 / 100,000 |
| Stargardt disease | 7 (2.1%) | 1-5 / 10,000 |

**Figure 6.**
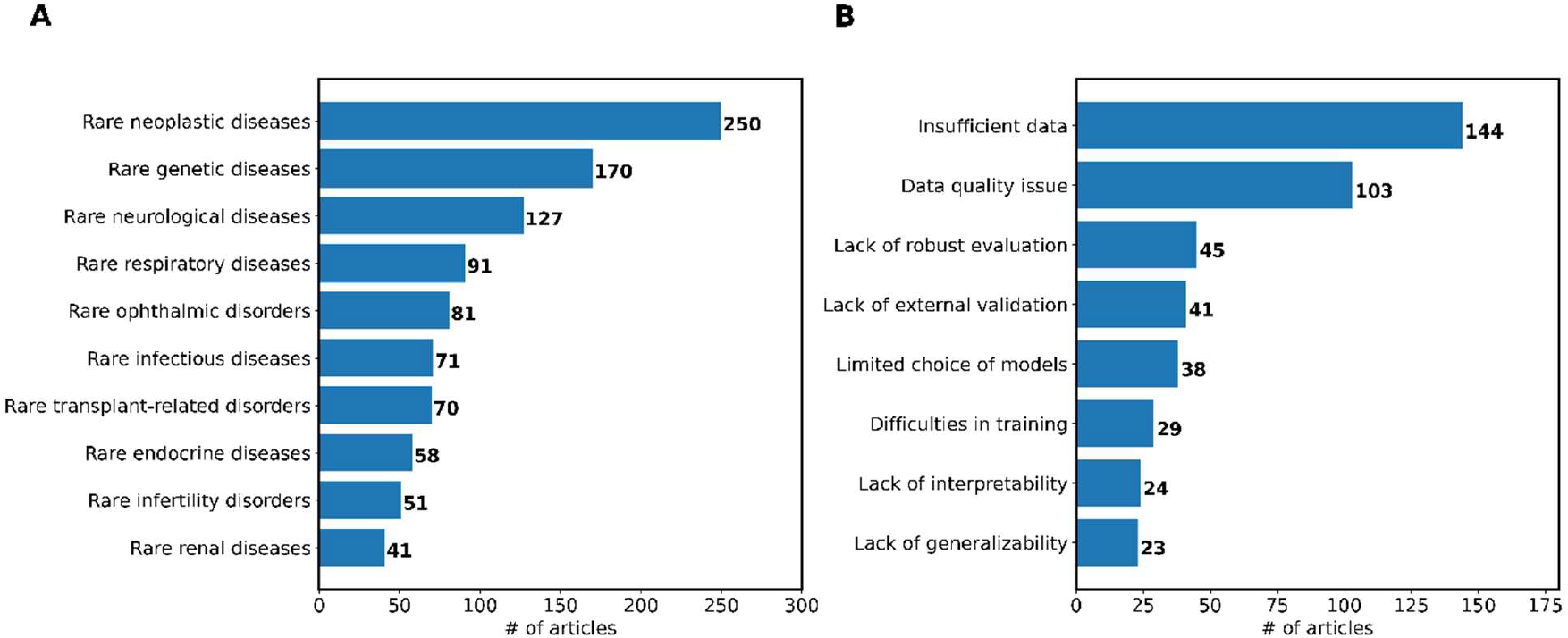
**(A)** The 10 most frequently studied rare disease groups in the reviewed articles. (**B**) The number of articles identified for each challenge.

### Challenges in Using Deep Learning for Rare Diseases

We classified the challenges in using deep learning for rare disease research reported by the authors into eight categories. Insufficient data is the most common challenge in the reviewed articles and were reported by 144 articles (43.4%). The size of the data used in the studies was relatively small compared to studies in other domains. For instance, the median size of image data used in the studies was 3,843 while the size of commonly used datasets for image classification task was from tens of thousands to tens of millions[50, 87]. 104 articles (31.3%) claimed that enhancing data quality can improve the performance of analytic task but come at the expense of manual examination and annotation of the data by experts (e.g., addressing selection bias or class imbalance of data), which is difficult to scale.

45 articles (13.6%) reported the lack of robust evaluation, primarily due to small size of data for evaluation and lack of clinically meaningful evaluation. Many studies attempted to overcome this challenge by using data augmentation and publicly available datasets. The increasingly available shared data can potentially mitigate this challenge. The need for external validation of a deep learning model was also frequently noted (41 articles [12.3%]). External validation of a deep learning model is often based on publicly available benchmark dataset. However, constrained by patient privacy concerns, developing publicly sharable healthcare datasets is difficult, unlike other domains to which deep learning is commonly applied such as natural language processing or computer vision. These restrictions limit evaluation of studies using external data. 38 articles (11.4%) pointed out that the studies can improve by including more state-of-the-art deep learning architectures. 29 articles (8.7%) reported difficulties in training deep learning models. 23 articles (6.9%) were concerned with models’ limited generalizability to other rare diseases.

Improving interpretability is a pressing need in using deep learning to healthcare and medicine[88, 89]. While many studies achieved good performance on their target analytic tasks, 24 articles (7.2%) claimed that the studies need better interpretability of the model. Few studies attempted to improve interpretability using various methods such as Grad-CAM [90] and SHAP[91] but through understanding of how the model(s) are converting inputs to outputs in order to understand how they are achieving their analytic task is still restricted[92-98].

## Discussion

### “Architecture-data type” conjugates

Our analysis showed that CNNs were the most frequently used deep learning architecture, and image data were the most frequently used data type. We observed that most of the studies used CNNs with image data, which shows rare disease research using deep learning has been driven by the conjugate of CNNs-image data. This is primarily because CNNs have gained more attention than other deep learning architectures resulting in sufficient available data and readily accessible source codes to implement the deep learning architectures. Image data in healthcare (e.g., CT and MRI) are often generated with a standardized protocol and can be de-identified, thus making data available for rare disease research using deep learning. As a result, some rare diseases where medical imaging is more commonly used for diagnosis or prognosis have been more frequently investigated using deep learning: tuberculosis research using chest X-ray and malaria research using blood smear image. This “deep learning architecture-data type” conjugate was also observed in other deep learning architectures. AEs were often used with measurement data, particularly with omics data. AEs were used to transform high-dimensional omics data into low-dimensional representations containing useful information. Encounter data were generally used with RNNs due to their longitudinal nature.

### To overcome difficulties underlying rare disease research using deep learning

Most studies applied the off-the-shelf deep learning architectures to their analytic tasks without modifications of the architectures in consideration of the characteristics of the target analytic task or data type. We found that more than half of the articles used deep learning as proof-of-concept or without clarifying a purpose. This perhaps indicates that current rare disease research using deep learning has been driven by simply adopting deep learning architectures that showed successful performance in other application domains such as computer vision or natural language processing.

Deep learning was used to reduce manual effort for the target analytic task and to address lack of data issue in 65 and 31 articles respectively. These two purposes are worth emphasis since they are closely related to the underlying difficulties of rare disease research. Reducing manual effort for rare disease analytic task is important since rare disease research and its clinical application often suffers from the high cost of obtaining resources. For example, timely and accurate diagnosis of a rare disease in low-resource environments such as developing countries is difficult since large amount of time and resources of experts is required.

Lack of data is one of the biggest hurdles for rare disease research using deep learning, as emphasized in the analysis of challenges in using deep learning for rare diseases. In the articles with the purposes to mitigate the lack of data issue, diverse approaches with deep learning were used to achieve this goal. Several articles attempted to develop novel deep learning architecture for their analytic tasks under the lack of data condition[57, 73, 99-105]. One of the frequently used strategies for addressing the scarce data problem is data augmentation, by either modifying the original data[92, 106-110], or generating synthetic data[62, 64-67, 111]. Transfer learning after pre-training of the deep learning models on other large datasets was also used to mitigate the lack of data issue[112-116].

### Future directions

First, the amount of data is often insufficient to fully explore the potential of deep learning for rare diseases. While insufficient data was the most frequently identified challenge in the reviewed articles, only about 10% of the entire articles sought to resolve the challenge in their studies. Additionally, only a few studies among those articles leveraged deep learning to tackle the challenge of insufficient data rather than simple data augmentation using the existing data. We expect to see more studies tackle this challenge using deep learning, which will be beneficial for improving rare disease research.

Second, the current evaluation of deep learning architectures on analytic tasks for rare diseases is limited. More robust evaluations should be conducted with external validation and the broad inclusion of deep learning architectures. However, external validation requires data sharing. One approach to overcome barriers in data sharing is to use a common data model (CDM). While there are several available CDMs (e.g., Observational Medical Outcomes Partnership CDM[117]), existing CDMs were developed for common diseases, thus their compatibility with rare diseases needs to be investigated. Inclusion of more deep learning architectures for evaluation requires additional efforts. A benchmark study that includes comparison of various deep learning architectures on a specific analytic task will be beneficial for providing robust evaluation.

Third, current deep learning models that have been used in rare disease research lack interpretability. Improving interpretability has been considered particularly important in using deep learning to research in healthcare and medicine[88, 89] and has been noted in the deep learning community. Since lack of interpretability makes it difficult for clinicians and healthcare providers to understand and adopt deep learning, the lack of interpretability should be addressed.

While application of deep learning to rare diseases in the real-world clinical setting is an important advance, none of the reviewed articles sought to apply deep learning in the real-world clinical setting. This is perhaps because of additional challenges to the aforementioned gaps to utilize deep learning for rare disease research. For example, a single institution or hospital only can see a small number of rare disease patients, which makes it challenging to apply deep learning for rare diseases. Another challenge is that deep learning models often require large amount of time for training and for conducting the analytic task. This can be a critical challenge in the real-world clinical application, where obtaining results of the analytic task in a timely and swift manner is important. Although there exist many challenges, we believe the field will benefit from studies that attempt to perform real-world application of deep learning for rare diseases.

### Limitations

There are some limitations to our study. First, identification and classification of rare diseases was solely based on Orphanet, thus there could be discrepancies based on other rare disease knowledge sources even though Orphanet provided monthly-updated rare disease definitions based on published scientific literature and expert reviews. For example, infectious diseases such as tuberculosis and malaria are often not recognized as rare diseases but we included them based on the Orphanet. Second, since our analysis did not evaluate the performance of deep learning architectures on their analytic tasks, our findings cannot be used to evaluate or compare different deep learning architectures on a specific analytic task.

## Conclusion

In this study, we conducted a scoping review of deep learning for rare disease studies with a focus on deep learning architecture. We found that convolutional neural networks were predominantly used for rare disease research utilizing image data. Future work can include (1) investigation about which rare diseases are best studied by which deep learning architecture(s); (2) how deep learning can further address the challenges underlying rare disease research; and (3) how deep learning can be applied in real-world practice for rare disease.

## Data Availability

All data produced in the present study are available upon reasonable request to the authors.

## Acknowledgements

This work was supported by National Institutes of Health grant R01LM012895.

## Data Availability

All data supporting the findings of this study are available in the paper and its supplementary material.

## Author Contributions

JL contributed to conceptualization of the study, literature search, abstract screening, full-text screening, data extraction, and data analysis. JK, ZC, and YS contributed to abstract screening, full text screening, and data extraction. CL contributed to conceptualization of the study and data analysis. JRR and WKC contributed to data analysis. CW supervised the study and contributed to conceptualization of the study and data analysis. All authors contributed to manuscript drafting and revision.

## REFERENCES

1. Vickers PJ. Challenges and opportunities in the treatment of rare diseases. Drug Discov World 2013;14:9–16

2. Wakap SN, Lambert DM, Olry A, et al. Estimating cumulative point prevalence of rare diseases: analysis of the Orphanet database. European Journal of Human Genetics 2020;28(2):165–73

3. Ronicke S, Hirsch MC, Türk E, Larionov K, Tientcheu D, Wagner AD. Can a decision support system accelerate rare disease diagnosis? Evaluating the potential impact of Ada DX in a retrospective study. Orphanet journal of rare diseases 2019;14(1):1–12

4. Schaefer J, Lehne M, Schepers J, Prasser F, Thun S. The use of machine learning in rare diseases: a scoping review. Orphanet Journal of Rare Diseases 2020;15(1):1–10

5. LeCun Y, Bengio Y, Hinton G. Deep learning. nature 2015;521(7553):436–44

6. Topol E. Deep medicine: how artificial intelligence can make healthcare human again: Hachette UK, 2019.

7. Rajkomar A, Dean J, Kohane I. Machine learning in medicine. N Engl J Med 2019;380(14):1347–58

8. Doctor ai: Predicting clinical events via recurrent neural networks. Machine Learning for Healthcare Conference; 2016.

9. Dipole: Diagnosis prediction in healthcare via attention-based bidirectional recurrent neural networks. Proceedings of the 23rd ACM SIGKDD international conference on knowledge discovery and data mining; 2017.

10. Choi E, Schuetz A, Stewart WF, Sun J. Using recurrent neural network models for early detection of heart failure onset. J Am Med Inform Assoc 2017;24(2):361–70

11. Lee J, Ta C, Kim JH, Liu C, Weng C. Severity Prediction for COVID-19 Patients via Recurrent Neural Networks. medRxiv 2020

12. Rasmy L, Xiang Y, Xie Z, Tao C, Zhi D. Med-BERT: pretrained contextualized embeddings on large-scale structured electronic health records for disease prediction. NPJ digital medicine 2021;4(1):1–13

13. De Freitas JK, Johnson KW, Golden E, et al. Phe2vec: Automated Disease Phenotyping based on Unsupervised Embeddings from Electronic Health Records. medRxiv 2021:2020.11. 14.20231894

14. Rifaioglu AS, Atas H, Martin MJ, Cetin-Atalay R, Atalay V, Doğan T. Recent applications of deep learning and machine intelligence on in silico drug discovery: methods, tools and databases. Briefings in bioinformatics 2019;20(5):1878–912

15. Tricco AC, Lillie E, Zarin W, et al. PRISMA extension for scoping reviews (PRISMA-ScR): checklist and explanation. Annals of internal medicine 2018;169(7):467–73

16. Orphanet: the portal for rare diseases and orphan drugs. Secondary Orphanet: the portal for rare diseases and orphan drugs. https://www.orpha.net.

17. Covidence. Secondary Covidence. https://www.covidence.org/.

18. Bakas S, Reyes M, Jakab A, et al. Identifying the best machine learning algorithms for brain tumor segmentation, progression assessment, and overall survival prediction in the BRATS challenge. arXiv preprint 1811.02629 2018

19. Johnson AE, Pollard TJ, Shen L, et al. MIMIC-III, a freely accessible critical care database. Sci. Data 2016;3(1):1–9

20. The Cancer Genome Atlas Program. Secondary The Cancer Genome Atlas Program. https://www.cancer.gov/about-nci/organization/ccg/research/structural-genomics/tcga.

21. Goodfellow I, Bengio Y, Courville A. Deep learning: MIT press, 2016.

22. Maggio V, Chierici M, Jurman G, Furlanello C. Distillation of the clinical algorithm improves prognosis by multi-task deep learning in high-risk neuroblastoma. PloS one 2018;13(12):e0208924

23. Fuhad K, Tuba JF, Sarker M, et al. Deep learning based automatic malaria parasite detection from blood smear and its smartphone based application. Diagnostics 2020;10(5):329

24. Chassagnon G, Vakalopoulou M, Régent A, et al. Deep learning–based approach for automated assessment of interstitial lung disease in systemic sclerosis on CT images. Radiology: Artificial Intelligence 2020;2(4):e190006

25. Gu T, Zhao X. Integrating multi-platform genomic datasets for kidney renal clear cell carcinoma subtyping using stacked denoising autoencoders. Scientific reports 2019;9(1):1–11

26. Francescatto M, Chierici M, Rezvan Dezfooli S, Zandonà A, Jurman G, Furlanello C. Multi-omics integration for neuroblastoma clinical endpoint prediction. Biology direct 2018;13(1):1–12

27. Yang Y, Walker TM, Walker AS, et al. DeepAMR for predicting co-occurrent resistance of Mycobacterium tuberculosis. Bioinformatics 2019;35(18):3240–49

28. Zhang L, Lv C, Jin Y, et al. Deep learning-based multi-omics data integration reveals two prognostic subtypes in high-risk neuroblastoma. Frontiers in genetics 2018;9:477

29. Guo L-Y, Wu A-H, Wang Y-x, Zhang L-p, Chai H, Liang X-F. Deep learning-based ovarian cancer subtypes identification using multi-omics data. BioData Mining 2020;13(1):1–12

30. U-net: Convolutional networks for biomedical image segmentation. International Conference on Medical image computing and computer-assisted intervention; 2015. Springer.

31. V-net: Fully convolutional neural networks for volumetric medical image segmentation. 2016 fourth international conference on 3D vision (3DV); 2016. IEEE.

32. Diller GP, Orwat S, Vahle J, et al. Prediction of prognosis in patients with tetralogy of Fallot based on deep learning imaging analysis. Heart 2020;106(13):1007–14

33. Kingma DP, Welling M. Auto-encoding variational bayes. arXiv preprint 1312.6114 2013

34. Chen X, Zeng M, Tong Y, et al. Automatic prediction of MGMT status in glioblastoma via deep learning-based MR image analysis. BioMed Research International 2020;2020

35. Romero R, Ramanathan A, Yuen T, et al. Mechanism of glucocerebrosidase activation and dysfunction in Gaucher disease unraveled by molecular dynamics and deep learning. Proceedings of the National Academy of Sciences 2019;116(11):5086–95

36. Simonyan K, Zisserman A. Very deep convolutional networks for large-scale image recognition. arXiv preprint 1409.1556 2014

37. Going deeper with convolutions. Proceedings of the IEEE conference on computer vision and pattern recognition; 2015.

38. Deep residual learning for image recognition. Proceedings of the IEEE conference on computer vision and pattern recognition; 2016.

39. Rethinking the inception architecture for computer vision. Proceedings of the IEEE conference on computer vision and pattern recognition; 2016.

40. Densely connected convolutional networks. Proceedings of the IEEE conference on computer vision and pattern recognition; 2017.

41. A Study on Tuberculosis Classification in Chest X-ray Using Deep Residual Attention Networks. 2020 42nd Annual International Conference of the IEEE Engineering in Medicine & Biology Society (EMBC); 2020. IEEE.

42. Arsalan M, Baek NR, Owais M, Mahmood T, Park KR. Deep Learning-Based detection of pigment signs for analysis and diagnosis of retinitis pigmentosa. Sensors 2020;20(12):3454

43. Kamnitsas K, Ledig C, Newcombe VF, et al. Efficient multi-scale 3D CNN with fully connected CRF for accurate brain lesion segmentation. Medical image analysis 2017;36:61–78

44. Eijgelaar RS, Visser M, Müller DM, et al. Robust Deep Learning–based Segmentation of Glioblastoma on Routine Clinical MRI Scans Using Sparsified Training. Radiology: Artificial Intelligence 2020;2(5):e190103

45. Wan Y, Rahmat R, Price SJ. Deep learning for glioblastoma segmentation using preoperative magnetic resonance imaging identifies volumetric features associated with survival. Acta neurochirurgica 2020;162(12):3067–80

46. Rahmat R, Saednia K, Khani MRHH, Rahmati M, Jena R, Price SJ. Multi-scale segmentation in GBM treatment using diffusion tensor imaging. Computers in Biology and Medicine 2020;123:103815

47. Pennig L, Hoyer UCI, Goertz L, et al. Primary central nervous system lymphoma: clinical evaluation of automated segmentation on multiparametric MRI using deep learning. Journal of Magnetic Resonance Imaging 2021;53(1):259–68

48. Laukamp KR, Thiele F, Shakirin G, et al. Fully automated detection and segmentation of meningiomas using deep learning on routine multiparametric MRI. European radiology 2019;29(1):124–32

49. Laukamp KR, Pennig L, Thiele F, et al. Automated meningioma segmentation in multiparametric MRI. Clinical neuroradiology 2021;31(2):357–66

50. Imagenet: A large-scale hierarchical image database. 2009 IEEE conference on computer vision and pattern recognition; 2009. Ieee.

51. Microsoft coco: Common objects in context. European conference on computer vision; 2014. Springer.

52. Calabrese E, Villanueva-Meyer JE, Cha S. A fully automated artificial intelligence method for non-invasive, imaging-based identification of genetic alterations in glioblastomas. Scientific reports 2020;10(1):1–11

53. Kriegsmann M, Haag C, Weis C-A, et al. Deep learning for the classification of small-cell and non-small-cell lung cancer. Cancers 2020;12(6):1604

54. Feng B, Hoskins W, Zhang Y, et al. Bi-stream CNN Down Syndrome screening model based on genotyping array. BMC medical genomics 2018;11(5):25–33

55. Yin B, Balvert M, van der Spek RA, et al. Using the structure of genome data in the design of deep neural networks for predicting amyotrophic lateral sclerosis from genotype. Bioinformatics 2019;35(14):i538–i47

56. Sengur A, Akbulut Y, Guo Y, Bajaj V. Classification of amyotrophic lateral sclerosis disease based on convolutional neural network and reinforcement sample learning algorithm. Health information science and systems 2017;5(1):1–7

57. Vance C, Kim Y, Zhang G, et al. Learning to detect the onset of slow activity after a generalized tonic–clonic seizure. BMC Medical Informatics and Decision Making 2020;20(12):1–8

58. Fabregat H, Araujo L, Martinez-Romo J. Deep neural models for extracting entities and relationships in the new RDD corpus relating disabilities and rare diseases. Computer methods and programs in biomedicine 2018;164:121–29

59. Enrich Rare Disease Phenotypic Characterizations via a Graph Convolutional Network Based Recommendation System. 2020 IEEE 33rd International Symposium on Computer-Based Medical Systems (CBMS); 2020. IEEE.

60. Kipf TN, Welling M. Semi-supervised classification with graph convolutional networks. arXiv preprint 1609.02907 2016

61. Keshavarzi Arshadi A, Salem M, Collins J, Yuan JS, Chakrabarti D. DeepMalaria: artificial intelligence driven discovery of potent antiplasmodials. Frontiers in pharmacology 2020;10:1526

62. Few-shot Radiology Report Generation for Rare Diseases. 2020 IEEE International Conference on Bioinformatics and Biomedicine (BIBM); 2020. IEEE.

63. Goodfellow I, Pouget-Abadie J, Mirza M, et al. Generative adversarial nets. Advances in neural information processing systems 2014;27

64. Rare Disease Prediction by Generating Quality-Assured Electronic Health Records∗. Proceedings of the 2020 SIAM International Conference on Data Mining; 2020. SIAM.

65. Li W, Wang Y, Cai Y, Arnold C, Zhao E, Yuan Y. Semi-supervised rare disease detection using generative adversarial network. arXiv preprint 1812.00547 2018

66. Yu K, Wang Y, Cai Y, et al. Rare disease detection by sequence modeling with generative adversarial networks. arXiv preprint 1907.01022 2019

67. Conan: Complementary pattern augmentation for rare disease detection. Proceedings of the AAAI Conference on Artificial Intelligence; 2020.

68. Tie X, Lam SK, Zhang Y, Lee KH, Au KH, Cai J. Pseudo-CT generation from multi-parametric MRI using a novel multi-channel multi-path conditional generative adversarial network for nasopharyngeal carcinoma patients. Medical physics 2020;47(4):1750–62

69. Hu X, Guo R, Chen J, et al. Coarse-to-fine adversarial networks and zone-based uncertainty analysis for NK/T-cell lymphoma segmentation in CT/PET images. IEEE journal of biomedical and health informatics 2020;24(9):2599–608

70. Casella A, Moccia S, Frontoni E, Paladini D, De Momi E, Mattos LS. Inter-foetus membrane segmentation for TTTS using adversarial networks. Annals of biomedical engineering 2020;48(2):848–59

71. Torrents-Barrena J, López-Velazco R, Piella G, et al. TTTS-GPS: Patient-specific preoperative planning and simulation platform for twin-to-twin transfusion syndrome fetal surgery. Computer methods and programs in biomedicine 2019;179:104993

72. Hochreiter S, Schmidhuber J. Long short-term memory. Neural computation 1997;9(8):1735–80

73. Multi-task learning via adaptation to similar tasks for mortality prediction of diverse rare diseases. AMIA Annual Symposium Proceedings; 2020. American Medical Informatics Association.

74. Ryu SM, Seo SW, Lee S-H. Novel prognostication of patients with spinal and pelvic chondrosarcoma using deep survival neural networks. BMC medical informatics and decision making 2020;20(1):1–10

75. Han I, Kim JH, Park H, Kim H-S, Seo SW. Deep learning approach for survival prediction for patients with synovial sarcoma. Tumor Biology 2018;40(9):1010428318799264

76. Ceccarelli F, Sciandrone M, Perricone C, et al. Prediction of chronic damage in systemic lupus erythematosus by using machine-learning models. PLoS One 2017;12(3):e0174200

77. Jang B-S, Jeon SH, Kim IH, Kim IA. Prediction of pseudoprogression versus progression using machine learning algorithm in glioblastoma. Scientific reports 2018;8(1):1–9

78. MRI to MGMT: predicting methylation status in glioblastoma patients using convolutional recurrent neural networks. PACIFIC SYMPOSIUM ON BIOCOMPUTING 2018: Proceedings of the Pacific Symposium; 2018. World Scientific.

79. Davidson B, Kalitzeos A, Carroll J, et al. Automatic cone photoreceptor localisation in healthy and Stargardt afflicted retinas using deep learning. Scientific reports 2018;8(1):1–13

80. Graves A, Schmidhuber J. Framewise phoneme classification with bidirectional LSTM and other neural network architectures. Neural networks 2005;18(5-6):602–10

81. Attallah O. CoMB-deep: composite deep learning-based pipeline for classifying childhood medulloblastoma and its classes. Frontiers in neuroinformatics 2021;15:21

82. Vaswani A, Shazeer N, Parmar N, et al. Attention is all you need. arXiv preprint 1706.03762 2017

83. Devlin J, Chang M-W, Lee K, Toutanova K. Bert: Pre-training of deep bidirectional transformers for language understanding. arXiv preprint 1810.04805 2018

84. Radiology Report Generation for Rare Diseases via Few-shot Transformer. 2021 IEEE International Conference on Bioinformatics and Biomedicine (BIBM); 2021. IEEE.

85. Prakash PKS, Chilukuri S, Ranade N, Viswanathan S. RareBERT: Transformer Architecture for Rare Disease Patient Identification using Administrative Claims. Thirty-Fifth AAAI Conference on Artificial Intelligence, AAAI 2021, Thirty-Third Conference on Innovative Applications of Artificial Intelligence, IAAI 2021, The Eleventh Symposium on Educational Advances in Artificial Intelligence, EAAI 2021, Virtual Event, February 2-9, 2021: AAAI Press, 2021:453–60.

86. Rare Disease Identification from Clinical Notes with Ontologies and Weak Supervision. 2021 43rd Annual International Conference of the IEEE Engineering in Medicine & Biology Society (EMBC); 2021 1–5 Nov. 2021.

87. Krizhevsky A, Hinton G. Learning multiple layers of features from tiny images. 2009

88. Interpretable machine learning in healthcare. Proceedings of the 2018 ACM international conference on bioinformatics, computational biology, and health informatics; 2018.

89. Vellido A. The importance of interpretability and visualization in machine learning for applications in medicine and health care. Neural computing and applications 2020;32(24):18069–83

90. Grad-cam: Visual explanations from deep networks via gradient-based localization. Proceedings of the IEEE international conference on computer vision; 2017.

91. Lundberg SM, Lee S-I. A unified approach to interpreting model predictions. Advances in neural information processing systems 2017;30

92. Sánchez Fernández I, Yang E, Calvachi P, et al. Deep learning in rare disease. Detection of tubers in tuberous sclerosis complex. PloS one 2020;15(4):e0232376

93. Yoon J, Han J, Park JI, et al. Optical coherence tomography-based deep-learning model for detecting central serous chorioretinopathy. Scientific reports 2020;10(1):1–9

94. Brown JM, Campbell JP, Beers A, et al. Automated diagnosis of plus disease in retinopathy of prematurity using deep convolutional neural networks. JAMA ophthalmology 2018;136(7):803–10

95. Wang S, Liu Z, Rong Y, et al. Deep learning provides a new computed tomography-based prognostic biomarker for recurrence prediction in high-grade serous ovarian cancer. Radiotherapy and Oncology 2019;132:171–77

96. Rajaraman S, Silamut K, Hossain MA, et al. Understanding the learned behavior of customized convolutional neural networks toward malaria parasite detection in thin blood smear images. Journal of Medical Imaging 2018;5(3):034501

97. Pasa F, Golkov V, Pfeiffer F, Cremers D, Pfeiffer D. Efficient deep network architectures for fast chest X-ray tuberculosis screening and visualization. Scientific reports 2019;9(1):1–9

98. Kubach J, Muhlebner-Fahrngruber A, Soylemezoglu F, et al. Same same but different: A Web-based deep learning application revealed classifying features for the histopathologic distinction of cortical malformations. Epilepsia 2020;61(3):421–32

99. ST-MetaDiagnosis: Meta learning with Spatial Transform for rare skin disease Diagnosis. 2020 IEEE International Conference on Bioinformatics and Biomedicine (BIBM); 2020. IEEE.

100. Difficulty-aware meta-learning for rare disease diagnosis. International Conference on Medical Image Computing and Computer-Assisted Intervention; 2020. Springer.

101. Deep multi-modality collaborative learning for distant metastases predication in PET-CT soft-tissue sarcoma studies. 2019 41st Annual International Conference of the IEEE Engineering in Medicine and Biology Society (EMBC); 2019. IEEE.

102. Kim T, Heo J, Jang D-K, et al. Machine learning for detecting moyamoya disease in plain skull radiography using a convolutional neural network. EBioMedicine 2019;40:636–42

103. Yang Q, Guo Y, Ou X, Wang J, Hu C. Automatic T staging using weakly supervised deep learning for nasopharyngeal carcinoma on MR images. Journal of Magnetic Resonance Imaging 2020;52(4):1074–82

104. Abdolmanafi A, Duong L, Dahdah N, Adib IR, Cheriet F. Characterization of coronary artery pathological formations from OCT imaging using deep learning. Biomedical optics express 2018;9(10):4936–60

105. Fu Y, Xue P, Ji H, Cui W, Dong E. Deep model with Siamese network for viable and necrotic tumor regions assessment in osteosarcoma. Medical Physics 2020;47(10):4895–905

106. Gao XW, Qian Y. Prediction of multidrug-resistant TB from CT pulmonary images based on deep learning techniques. Molecular pharmaceutics 2017;15(10):4326–35

107. Yun J, Park JE, Lee H, Ham S, Kim N, Kim HS. Radiomic features and multilayer perceptron network classifier: a robust MRI classification strategy for distinguishing glioblastoma from primary central nervous system lymphoma. Scientific reports 2019;9(1):1–10

108. Banerjee I, Crawley A, Bhethanabotla M, Daldrup-Link HE, Rubin DL. Transfer learning on fused multiparametric MR images for classifying histopathological subtypes of rhabdomyosarcoma. Computerized Medical Imaging and Graphics 2018;65:167–75

109. Prince EW, Whelan R, Mirsky DM, et al. Robust deep learning classification of adamantinomatous craniopharyngioma from limited preoperative radiographic images. Scientific reports 2020;10(1):1–13

110. Shah M, Roomans Ledo A, Rittscher J. Automated classification of normal and Stargardt disease optical coherence tomography images using deep learning. Acta ophthalmologica 2020;98(6):e715–e21

111. Using synthetic training data for deep learning-based GBM segmentation. 2019 41st Annual International Conference of the IEEE Engineering in Medicine and Biology Society (EMBC); 2019. IEEE.

112. How well do U-Net-based segmentation trained on adult cardiac magnetic resonance imaging data generalize to rare congenital heart diseases for surgical planning? Medical Imaging 2020: Image-Guided Procedures, Robotic Interventions, and Modeling; 2020. International Society for Optics and Photonics.

113. Detection of rare genetic diseases using facial 2D images with transfer learning. 2018 8th International Symposium on Embedded Computing and System Design (ISED); 2018. IEEE.

114. Ziegelmayer S, Kaissis G, Harder F, et al. Deep Convolutional Neural Network-Assisted Feature Extraction for Diagnostic Discrimination and Feature Visualization in Pancreatic Ductal Adenocarcinoma (PDAC) versus Autoimmune Pancreatitis (AIP). Journal of clinical medicine 2020;9(12):4013

115. Gudmundsson E, Straus CM, Li F, Armato SG. Deep learning-based segmentation of malignant pleural mesothelioma tumor on computed tomography scans: application to scans demonstrating pleural effusion. Journal of Medical Imaging 2020;7(1):012705

116. Lao J, Chen Y, Li Z-C, et al. A deep learning-based radiomics model for prediction of survival in glioblastoma multiforme. Scientific reports 2017;7(1):1–8

117. Hripcsak G, Duke JD, Shah NH, et al. Observational Health Data Sciences and Informatics (OHDSI): opportunities for observational researchers. Stud Health Technol Inform 2015;216:574

